# Textbook-Level Medical Knowledge in Large Language Models: A Comparative Evaluation Using the Japanese National Medical Examination

**DOI:** 10.1101/2025.09.10.25335398

**Authors:** Mingxin Liu, Tsuyoshi Okuhara, Zhehao Dai, Minghong Zhao, Wenqiang Yin, Hiroko Okada, Emi Furukawa, Takahiro Kiuchi

## Abstract

**Study aims and objectives:** This study aimed to evaluate the performance of four reasoning-enhanced large language models (LLMs)—GPT-5, Grok-4, Claude Opus 4.1, and Gemini 2.5 Pro—on the Japanese National Medical Examination (JNME).

**Methods:** We evaluated LLM performance using the 2019 and 2025 JNME (n = 793). Questions were entered into each model with chain-of-thought prompting enabled. Accuracy was assessed overall as well as by question type, content domain, and difficulty. Incorrect responses were qualitatively reviewed by a licensed physician and a medical student.

**Results:** From highest to lowest, the overall accuracies of the four LLMs were 97.2% for Gemini 2.5 Pro, 96.3% for GPT-5, 96.1% for Claude Opus 4.1, and 95.6% for Grok-4, with no significant pairwise differences observed. All four LLMs reached the threshold generally regarded as sufficient to serve as reliable medical knowledge sources. Question type (e.g., image-based, clinically oriented, and difficult items) still influenced LLM performance, but the performance gaps were much smaller than in earlier generations of LLMs. Notably, Gemini 2.5 Pro consistently achieved the highest performance, including 96.1% on image-based questions and 97.0% on clinical questions. Common error patterns included providing extra response options and misinterpreting laterality when analyzing X-ray images or computed tomography (CT).

**Conclusions:** Advanced LLMs released in 2025 achieved textbook-level accuracy on the JNME, surpassing the 95% benchmark for reliable knowledge sources. Gemini 2.5 Pro achieved the highest accuracy across all question types and demonstrated the greatest stability, while Grok-4 showed more variability. These findings highlight a milestone in which LLMs have achieved the level necessary to be considered educational resources and decision-support tools.

**Statements and Declarations:** This work was supported by JSPS KAKENHI Grant Number 24KJ0830.

## 1. Introduction

### 1.1. Background

Since OpenAI introduced ChatGPT in November 2022, the first widely adopted artificial intelligence (AI) chatbot powered by a large language model (LLM), these systems have rapidly attracted worldwide interest because of their ability to generate elaborate responses to sophisticated questions [1]. Within the medical domain, LLMs are increasingly recognized for their potential contributions to both clinical decision-making and medical education [2–6]. Many LLMs incorporate image interpretation capabilities, enabling applications in areas such as dermatology and radiology, where they can assist in analyzing skin lesions or X-ray images [2,3]. Moreover, unlike conventional search engines, which return a list of hyperlinks, LLM-based chatbots are designed to deliver direct and practical answers, thereby functioning as accessible knowledge resources [2].

Despite these advances, the reliability of medical knowledge embedded in LLMs remains a critical hurdle for their integration into education and clinical workflows. Previous research has emphasized that, to serve as dependable medical-education tools, their response accuracy should consistently surpass 95% [7]. Several studies from different countries have used national medical licensing examinations to benchmark LLM performance [8–18]. A systematic review reported that GPT-4 achieved an average accuracy of approximately 81% across multiple national licensing exams, sufficient to pass many of them but remained insufficient to be considered a reliable knowledge source [4].

Building on this body of work, our 2024 study demonstrated that GPT-4o reached an accuracy of 89.2% on the Japanese National Medical Licensing Examination (JNME) [18]. Similarly, a recent Chinese investigation revealed that DeepSeek-R1 achieved 92% accuracy on the China National Medical Licensing Examination [19]. Notably, the study also highlighted the effectiveness of chain-of-thought (CoT) prompting, in which the model was instructed to articulate intermediate reasoning steps before providing a final answer, leading to significant performance improvements [19]. Collectively, these findings suggest that the most advanced LLMs are approaching, although not yet achieving, the critical 95% accuracy threshold required for reliable educational and clinical applications.

In 2025, a new wave of LLMs equipped with reasoning-enhancement features was introduced. In July 2025, Google and xAI released Gemini 2.5 Pro and Grok-4, respectively, and in August, Anthropic and OpenAI released Claude Opus 4.1 and GPT-5, respectively [20–23]. These models, which incorporate CoT techniques, have drawn considerable attention regarding whether they can achieve sufficiently high accuracy to be regarded as reliable knowledge sources. Thus, this study aimed to evaluate the performance of GPT-5, Grok-4, Claude Opus 4.1, and Gemini 2.5 Pro on the JNME to further examine both their overall applicability to medical education in Japan and the differences among the latest LLMs.

### 1.2. Study Aims and Objectives

In this study, we employed the JNME to assess the capabilities of the four latest LLMs: GPT-5, Grok-4, Claude Opus 4.1, and Gemini 2.5 Pro. Our evaluation was designed to address the following key questions.

1. What levels of accuracy can these LLMs achieve on the JNME, and can any of them pass the exam or meet the 95% threshold?
2. How does performance differ between image-based and text-only questions?
3. Do the LLMs show varying accuracy on general versus clinical questions?
4. Is their performance influenced by the publication year of the exam questions?
5. To what extent does question difficulty affect accuracy?
6. What characteristics are present in the questions that the LLMs answer incorrectly?

By systematically investigating these aspects, we aim to clarify the strengths and limitations of the latest LLMs in solving medical examination problems. Furthermore, we highlight persistent challenges and propose directions for future model refinement, thereby contributing to the integration of LLMs into medical education and clinical practice.

## 2. Methods

### 2.1. Tested LLMs

As of August 2025, four LLMs represent the most advanced publicly available systems: GPT-5, Grok-4, Claude Opus 4.1, and Gemini 2.5 Pro. These models were selected for evaluation in this study [20–23].

### 2.2. Japanese National Medical Licensing Examination (JNME)

The JNME was first introduced in 1946 as a national licensing test for medical school graduates who completed six years of advanced training. Over time, the exam has undergone several revisions, and its current structure has remained unchanged since 2018. The JNME consists of 400 questions divided into six sections (A–F). Sections A, C, D, and F are designated as non-essential, each containing 75 questions, whereas sections B and E are considered essential, each including 50 questions. The scoring systems differ by section. In the essential sections (B and E), general knowledge questions are worth one point each, and clinical questions carry three points, with a minimum of 160 points required to pass. In the non-essential sections (A, C, D, and F), all questions are assigned one point, and the cutoff score is not predetermined but generally falls around 220 points. Additionally, the exam includes approximately 10 multiple-choice questions (MCQs) with “taboo” choices, where selecting more than three of these prohibited options automatically results in failure.

The average annual pass rate for Japanese medical students is approximately 90%. The question formats includes both MCQs and calculation-based items. MCQs appear in several formats: five-option single-answer, five-option multiple-answer (requiring two or three selections), and extended-option types with more than six options. For multiple-answer items, the number of correct responses is explicitly stated. The exam also integrates image-based questions to assess visual diagnostic skills [24].

### 2.3. Questions Utilized in This Study

As our previous studies had already employed the 2018 and 2024 versions of the JNME, we avoided reusing them to minimize potential data contamination. For this analysis, we utilized the entire set of questions from the 2019 and 2025 JNME. This choice served two purposes: first, to ensure independence from previous work, and second, to allow a clear comparison of LLM performance on exam items created before and after the models’ training cutoff dates. Specifically, the Ministry of Health, Labour, and Welfare of Japan released the official questions and answers to the 2025 JNME on April 28, 2025 [25]. The knowledge cutoff dates for the four evaluated models were September 2024 for GPT-5 [26], November 2024 for Grok-4 [27], January 2025 for Gemini 2.5 Pro [28], and March 2025 for Claude Opus 4.1 [29]. Consequently, none of the LLMs had prior access to the content of the 2025 JNME.

To facilitate a more detailed evaluation, we classified the exam questions based on the following criteria:

1. Question type: image-based versus non-image-based.
2. Content domain: general versus clinical questions.
3. Difficulty level: based on the answer statistics published by Medu4, a preparatory school for the JNME, items were categorized into three groups—easy (≥90% of medical students answered correctly), moderate (70–89%), and difficult (<70%).

### 2.4. Inputting Questions to LLMs

We input these questions into the LLMs between August 8 and August 20, 2025. Both the textual content and images from the exam were entered directly into each LLM’s chat interface. To ensure that each model operated under its best reasoning settings, we activated reasoning enhancement where available: GPT-5 was tested using its “thinking mode” and Claude Opus 4.1 using the “extended thinking” option. Grok-4 and Gemini 2.5 Pro were evaluated in their default configurations, which incorporated internal reasoning enhancements. Internet search functions were disabled to minimize the risk of data contamination. For Grok-4, which does not provide a direct option to disable web search, we applied the following explicit instruction: “Disable Grok from performing any network searches when generating responses.”

Moreover, to avoid contextual interference from previous interactions, each examination item was presented in a new, independent chat. Exceptions were made only for sequential questions that required continuity, which were entered in the same chat. The order of the questions followed that of the original examination, and each question was presented once. If an LLM failed to generate a response owing to system issues, the item was resubmitted until a valid answer was produced.

No additional prompts were provided for answering these questions. However, in rare cases where a model declined to respond, we employed a clarification prompt—“This is a question from the medical licensing examination”—to elicit an answer. All responses were recorded in an Excel spreadsheet and two independent authors (MX Liu and MH Zhao) assessed each output as correct or incorrect.

### 2.5. Statistical Analysis

Descriptive statistics were calculated to summarize model performance, including the total number of questions, number of correct responses, accuracy proportions, and mean values. The accuracy rates across different LLMs and question categories were compared using Fisher’s exact test. For multiple comparisons, p-values were reported, with statistical significance set at p ≤ 0.05 (two-tailed). All analyses were performed using R software (version 4.4.0).

In addition to the quantitative analyses, all incorrect responses were reviewed by two co-authors: ZH Dai, a licensed physician in Japan, and WQ Yin, a medical student preparing for the JNME. The review examined both the explanations and reasoning traces provided by the LLMs, in order to identify common patterns of the incorrection.

### 2.6. Ethical Considerations

The JNME questions and LLMs used in this study were publicly accessible. Ethics approval was not required for this study.

## 3. Results

### 3.1. Characteristics of the JNME Questions

For the 2019 JNME dataset, four invalid questions and three questions containing non-public images were excluded, resulting in 393 questions available for analysis. All 400 questions from the 2025 JNME were retained. The combined dataset comprised 793 questions, of which 300 were classified as general and 493 as clinical. A total of 203 questions were image-based, and 590 were text-only. Based on difficulty levels, 437 questions were categorized as easy, 223 as moderate, and 133 as difficult.

### 3.2. Accuracy Rates of LLM Responses

All four LLMs successfully generated responses for the entire set of 793 questions. The complete outputs, along with their correctness annotations, are provided in supplementary materials 1. Of the 793 questions, GPT-5, Grok-4, Claude Opus 4.1, and Gemini 2.5 Pro correctly answered 764, 758, 762, and 771 questions, respectively. The corresponding overall accuracy rates, ranked from highest to lowest, were 97.2% for Gemini 2.5 Pro, 96.3% for GPT-5, 96.1% for Claude Opus 4.1, and 95.6% for Grok-4.

Pairwise comparisons revealed no statistically significant differences in accuracy among the four models (all p-values > 0.05). Similarly, for each LLM, the performance did not differ significantly between the 2025 and 2019 JNME (all p > 0.05) (Figure 1).

**Fig. 1.**
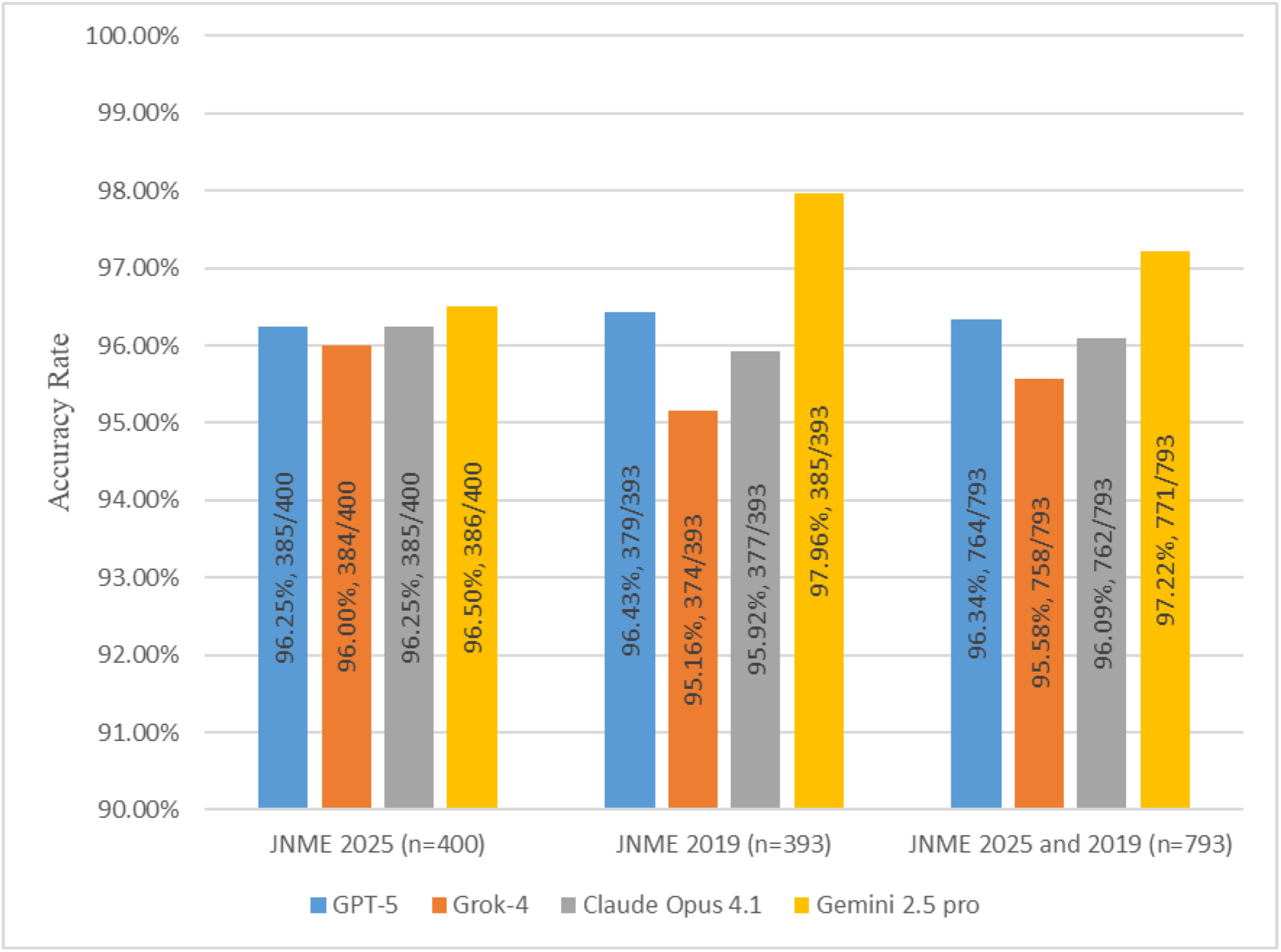
Overall correct number and accuracy of the four LLMs.

The examination scores of the four LLMs were calculated according to the official scoring rules of the JNME to determine whether they would pass the test. All four LLMs exceeded the passing line in both the 2019 and 2025 JNME, as well as in both essential and non-essential sections. Notably, for the essential sections, all four LLMs lost fewer than 10 points, achieving nearly perfect scores (Figure 2).

**Fig. 2.**
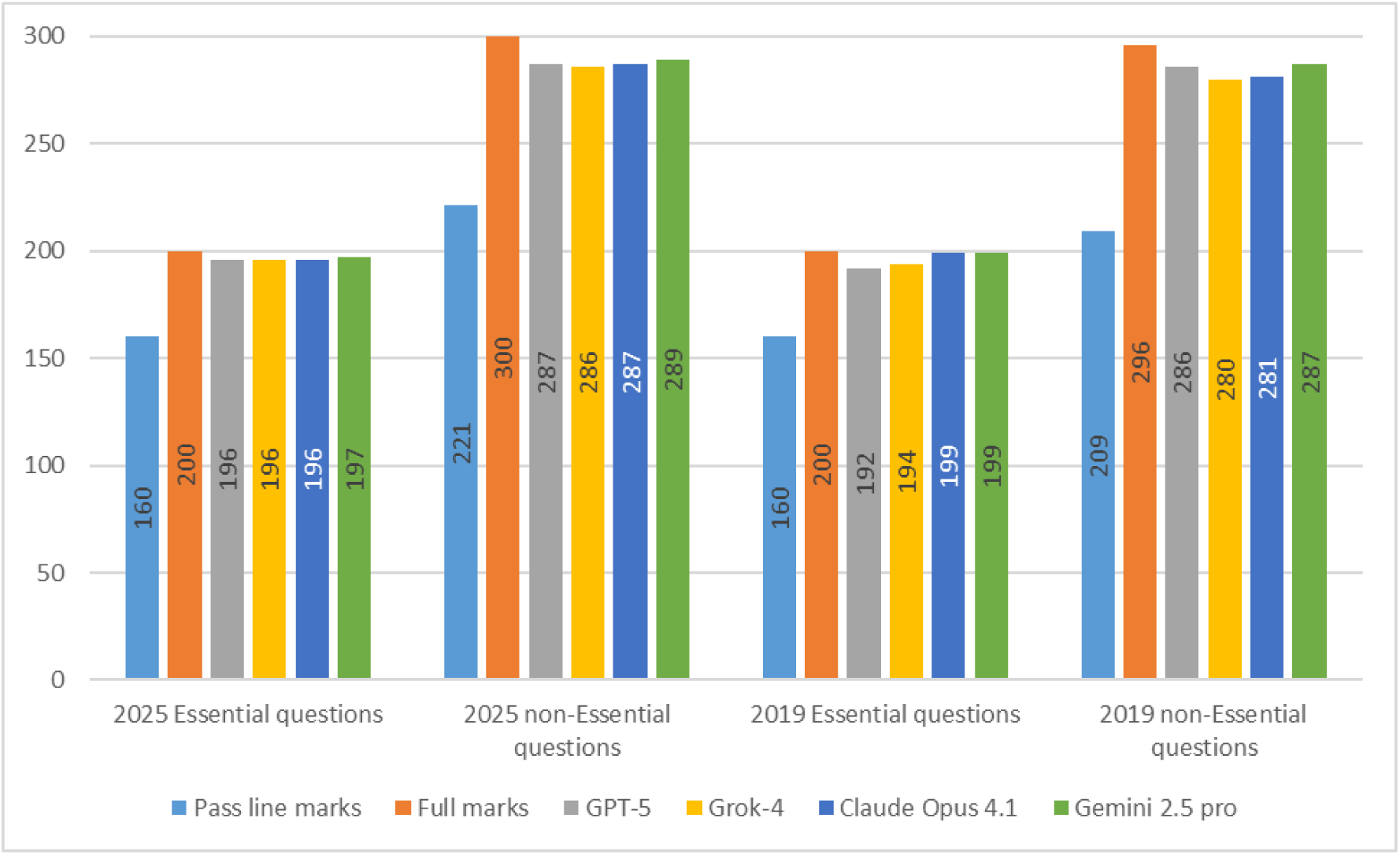
Score of each LLM calculated by the JNME scoring rules.

For non-image-based questions, GPT-5, Grok-4, Claude Opus 4.1, and Gemini 2.5 Pro achieved accuracies of 97.3%, 97.1%, 97.3%, and 97.6%, respectively, with no significant differences among the four LLMs (all p-values > 0.05). For image-based questions, the accuracies were 93.6%, 91.1%, 92.6%, and 96.1% for GPT-5, Grok-4, Claude Opus 4.1, and Gemini 2.5 Pro, respectively. A significant difference was observed only between Gemini 2.5 Pro and Grok-4 (p < 0.05) (Table 1). Across both the 2019 and 2025 JNME, the accuracies of all four LLMs were consistently higher for non-image-based questions than for image-based questions. The differences in accuracy rates between the two question types were 3.7%, 6.0%, 4.7%, and 1.5% for GPT-5, Grok-4, Claude Opus 4.1, and Gemini 2.5 Pro, respectively. Three of the LLMs showed significant differences (GPT-5, p = 0.016; Grok-4, p < 0.001; Claude Opus 4.1, p = 0.03) (Figure 3).

**Table 1.** Accuracy of image-based and non image-based questions.

|  |  | GPT-5 |  | Grok-4 |  | Claude Opus 4.1 |  | Gemini 2.5 pro |  |
| --- | --- | --- | --- | --- | --- | --- | --- | --- | --- |
|  |  | Correct number | Correct rate | Correct number | Correct rate | Correct number | Correct rate | Correct number | Correct rate |
| Questions with image | JNME 2025 (n=102) | 96 | 94.1% | 93 | 91.2% | 96 | 94.1% | 98 | 96.1% |
|  | JNME 2019 (n=101) | 94 | 93.1% | 92 | 91.1% | 92 | 91.1% | 97 | 96.0% |
|  | JNME 2025 and 2019 (n=203) | 190 | 93.6% | 185 | 91.1% | 188 | 92.6% | 195 | 96.1% |
| Questions without image | JNME 2025 (n=298) | 289 | 97.0% | 291 | 97.7% | 289 | 97.0% | 288 | 96.6% |
|  | JNME 2019 (n=292) | 285 | 97.6% | 282 | 96.6% | 285 | 97.6% | 288 | 98.6% |
|  | JNME 2025 and 2019 (n=590) | 574 | 97.3% | 573 | 97.1% | 574 | 97.3% | 576 | 97.6% |

**Fig 3.**
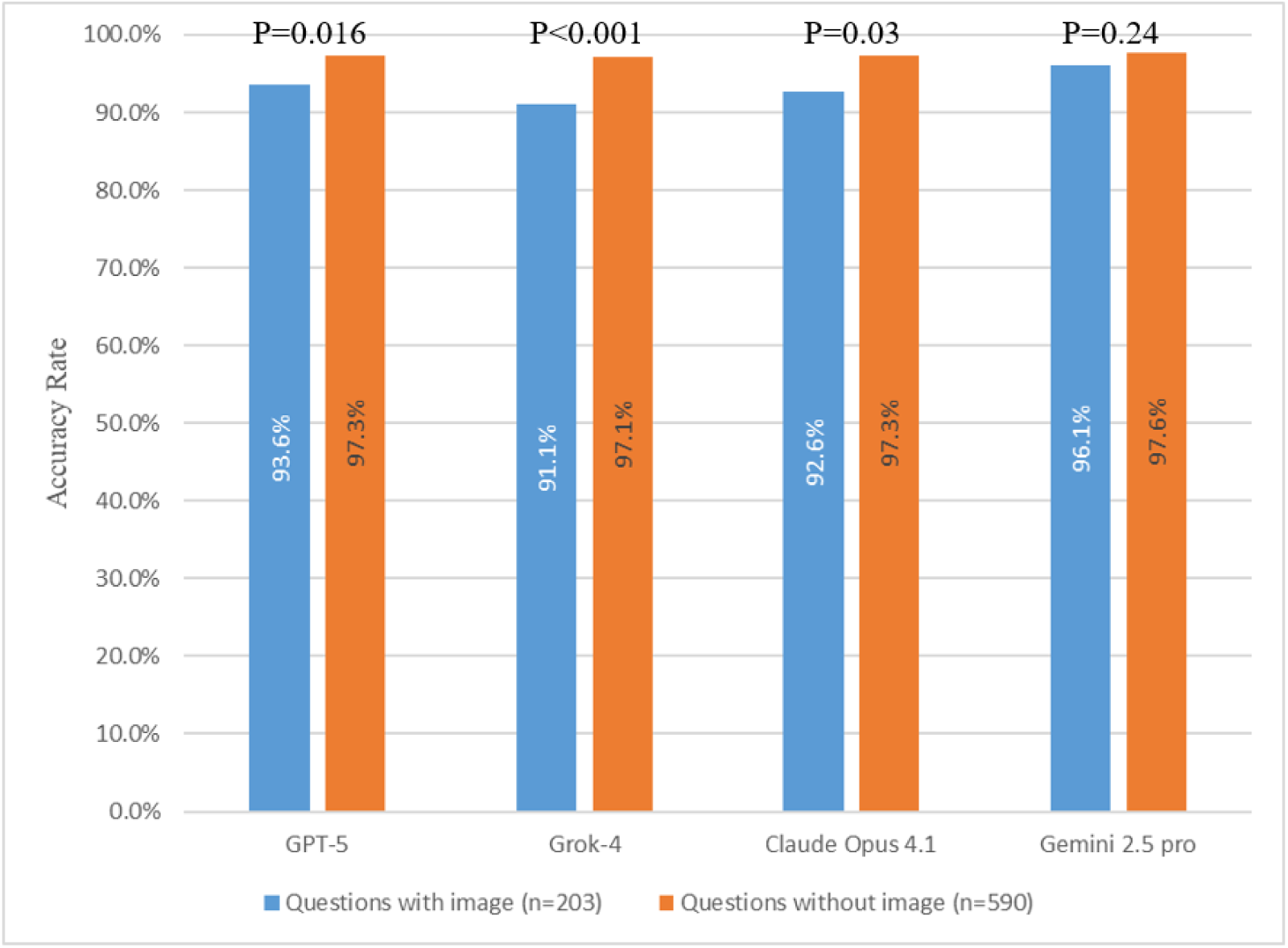
Accuracy of image-based and non-image-based questions.

For performance across different difficulty levels, the accuracy rates of GPT-5 were 97.5%, 96.4%, and 92.5% for easy, moderate, and difficult questions, respectively. Grok-4 achieved 98.2%, 93.7%, and 90.2% accuracy for easy, moderate, and difficult questions, respectively. Claude Opus 4.1 demonstrated accuracies of 98.2%, 97.3%, and 87.2%, whereas Gemini 2.5 Pro reached 98.4%, 97.3%, and 93.2% across the three difficulty levels (Table 2). When comparing easy versus difficult questions, all four LLMs showed significant differences (all p-values < 0.05) (Table 3). In comparisons between the easy and moderate questions, only Grok-4 showed a significant difference (p < 0.01) (Table 3). In comparisons between moderate and difficult questions, only Claude Opus 4.1 showed a significant difference (p < 0.001) (Table 3).

**Table 2.** Accuracy rate of each LLM on different difficulty level questions.

| GPT-5 | Grok-4 | Claude Opus 4.1 | Gemini 2.5 pro |
| --- | --- | --- | --- |

|  |  | Correct<br>number | Correct<br>rate | Correct<br>number | Correct<br>rate | Correct<br>number | Correct<br>rate | Correct<br>number | Correct<br>rate |
| --- | --- | --- | --- | --- | --- | --- | --- | --- | --- |
| **JNME**<br>**2025**<br>**(n=400)** | easy<br>(n=238,<br>59.5%) | 231 | 97.10% | 234 | 98.30% | 233 | 97.90% | 232 | 97.50% |
|  | moderate<br>(n=100,<br>25%) | 95 | 95.00% | 92 | 92.00% | 96 | 96.00% | 96 | 96.00% |
|  | difficult<br>(n=62,<br>15.5%) | 59 | 95.20% | 58 | 93.50% | 56 | 90.30% | 58 | 93.50% |
| **JNME**<br>**2019**<br>**(n=393)** | easy<br>(n=199,<br>50.6%) | 195 | 98.00% | 195 | 98.00% | 196 | 98.50% | 198 | 99.50% |
|  | moderate<br>(n=123,<br>31.3%) | 120 | 97.60% | 117 | 95.10% | 121 | 98.40% | 121 | 98.40% |
|  | difficult<br>(n=71,<br>18.1%) | 64 | 90.10% | 62 | 87.30% | 60 | 84.50% | 66 | 93.00% |
| **JNME**<br>**2025**<br>**and**<br>**2019**<br>**(n=793)** | easy<br>(n=437,<br>55.1%) | 426 | 97.50% | 429 | 98.20% | 429 | 98.20% | 430 | 98.40% |
|  | moderate<br>(n=223,<br>28.1%) | 215 | 96.40% | 209 | 93.70% | 217 | 97.30% | 217 | 97.30% |
|  | difficult<br>(n=133,<br>16.8%) | 123 | 92.50% | 120 | 90.20% | 116 | 87.20% | 124 | 93.20% |

**Table 3.** P-values for comparisons across difficulty levels of four LLMs.

|  | **GPT-5** | **Grok-4** | **Claude Opus 4.1** | **Gemini 2.5 pro** |
| --- | --- | --- | --- | --- |
| **Easy vs difficult** | p < 0.01 | p < 0.001 | p < 0.001 | p < 0.01 |
| **Easy vs moderate** | p = 0.437 | p < 0.01 | p = 0.468 | p = 0.341 |
| **Difficult vs moderate** | p = 0.101 | p = 0.228 | p < 0.001 | p = 0.064 |

For general questions, the accuracy rates of GPT-5, Grok-4, Claude Opus 4.1, and Gemini 2.5 Pro were 99.0%, 99.0%, 97.6%, and 98.6%, respectively. The corresponding accuracy rates for the clinical questions were 95.4%, 94.2%, 95.8%, and 97.0%, respectively (Table 4). All four LLMs demonstrated higher accuracy for general questions than for clinical questions; however, a statistically significant difference was observed only for Grok-4 (p < 0.001).

**Table 4.**
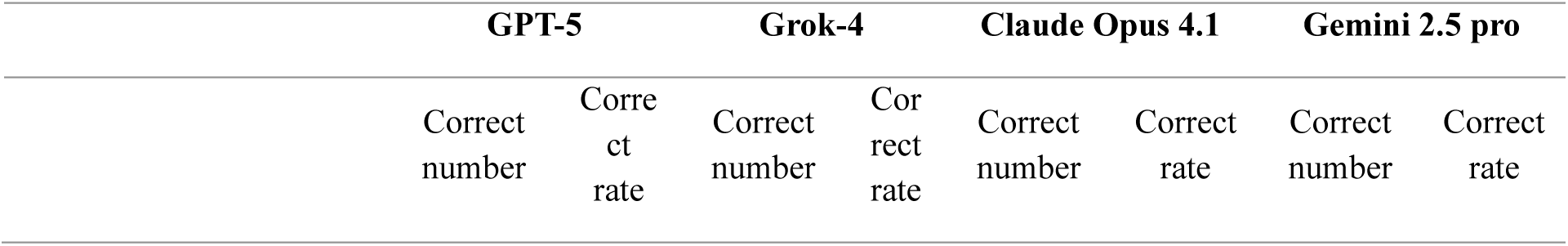
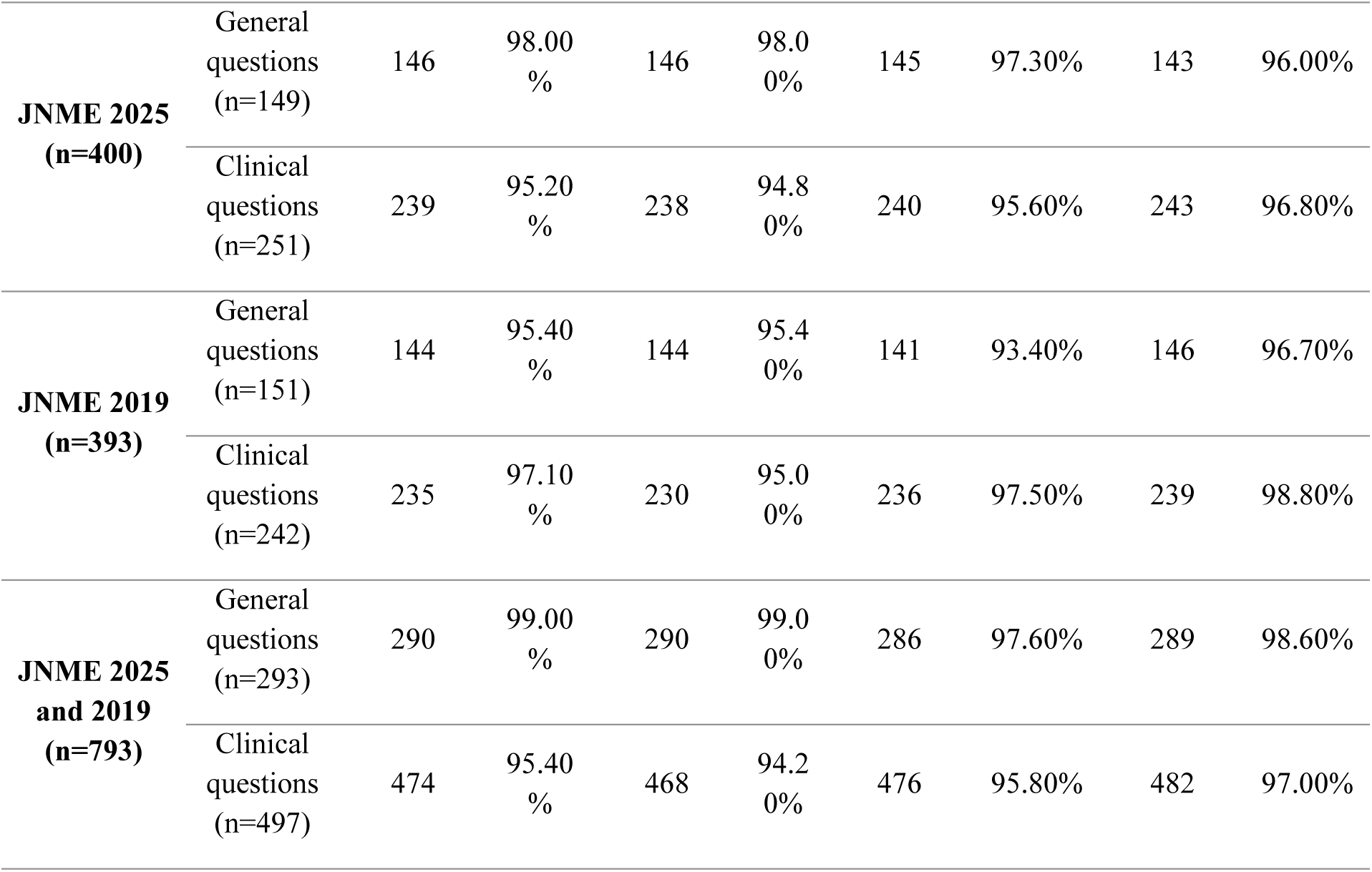
Accuracy rate of each LLM on general and clinical questions.

## 4. Discussion

### 4.1. Principal Findings

To the best of our knowledge, this is the first study to evaluate the performance of the most advanced reasoning-enhanced LLMs—GPT-5, Grok-4, Claude Opus 4.1, and Gemini 2.5 Pro—in medical licensing examinations.

According to the official JNME scoring rules, all four models passed both the 2019 and 2025 examinations, achieving near-perfect scores in the essential sections. None of the LLMs selected any taboo choices. In terms of overall accuracy (correct question numbers/total question numbers), the LLMs ranked as follows: Gemini 2.5 Pro (97.2%), GPT-5 (96.3%), Claude Opus 4.1 (96.1%), and Grok-4 (95.6%). All exceeded the 95% threshold, which has been proposed as a benchmark for considering LLMs as reliable sources of medical knowledge. This suggests that the medical knowledge encoded in these LLMs approaches textbook level, representing a milestone in their practical application as educational tools in medicine.

Notably, the accuracies achieved in this study are the highest reported to date among all evaluations of LLMs for medical licensing examinations worldwide, significantly surpassing the performance of GPT-4o (89.2% accuracy) in previous studies [18]. Furthermore, unlike earlier findings in which GPT-4o significantly outperformed Gemini 1.5 Pro and Claude 3 Opus [18], the present evaluation revealed no significant differences in accuracy among the four cutting-edge LLMs. We believe that these four pragmatic advances best explain this improvement over prior models. Firstly, reasoning enhancements that make models “think before answering”—including CoT, Self-Consistency, and search-style Tree-of-Thoughts—are known to improve performance on multistep problems typical of clinical reasoning and computation. Moreover, “process supervision” trains models to produce correct intermediate steps, not just correct final answers, which aligns well with medical exam questions requiring stepwise justification [19,30,31]. Second, the latest models plausibly benefit from broader and more recent corpora with improved coverage of medical content (e.g., guideline-like text and exam-style phrasing) and better Japanese biomedical material, narrowing the historical gap seen in earlier English-skewed systems [32]. Third, no instances of AI hallucinations were observed in any of the LLMs examined. Compared with earlier LLMs (e.g., GPT-4o and Gemini 1.5 pro), current LLMs more often ground their answers and self-check their drafts. Techniques like retrieval-augmented generation supply external evidence, while “chain-of-verification” style decoding has been shown to lower hallucinations by planning and answering verification questions before finalizing a response—both of which are valuable for factual, guideline-consistent medical Q&A [33].

All four LLMs achieved accuracies exceeding 97% for non-image-based questions, with Gemini 2.5 Pro demonstrating near-perfect performance by attaining the highest accuracy of 98.6%. Consistent with previous studies [18,34,35], their accuracies on image-based questions were lower than on non-image-based questions. GPT-5, Claude 4.1 Opus, and Grok 4 showed a statistically significant difference between image and non-image bases questions, whereas the Gemini 2.5 Pro did not. However, unlike previous studies, in which LLMs typically achieved less than 80% accuracy on image-based questions [4], the present study found that all four models exceeded 90% accuracy even on image-based questions, with Gemini 2.5 Pro reaching the highest accuracy of 96.1%. These findings suggest that the image interpretation capabilities of the latest generation of LLMs have substantially improved compared with earlier models. Notably, Gemini 2.5 Pro performed exceptionally well, achieving an accuracy above the 95% benchmark even for image-based questions, which supports its potential applicability as a medical imaging diagnostic tool. In contrast, the other three LLMs may pose risks if applied prematurely in medical imaging analyses or clinical diagnostic support.

We categorized all questions into three levels of difficulty (easy, moderate, and difficult) based on human medical students’ accuracy and evaluated the LLMs accordingly. All four models demonstrated significantly higher accuracy for easy questions than for difficult questions. However, no significant differences were observed between easy and moderate questions or between moderate and difficult questions. In the easy, moderate, and difficult categories, Gemini 2.5 Pro achieved the highest accuracy across the board. Importantly, except for Grok-4, the remaining three LLMs maintained accuracies above 90% even for difficult questions. These findings suggest that although previous studies have consistently reported that LLMs perform better on easier items [34,36–40], the performance gap attributable to difficulty narrows as LLM capabilities advance. Collectively, these patterns indicate enhanced stability across difficulty strata, stronger internal consistency in reasoning processes, and greater robustness to item complexity and distributional shifts. Consequently, the classical “difficulty effect” is attenuated, with model accuracies approaching a ceiling and variance across categories being substantially reduced. This enhanced uniformity across difficulty levels suggests that advanced LLMs may serve as more consistent and equitable tools for medical education and assessment, thereby reducing biases introduced by item complexity.

Typically, the performance of LLMs on general questions reflects their accuracy as knowledge sources in medical education, whereas their performance on clinical questions reflects their capabilities in clinical reasoning and diagnostic decision-making. By comparing the two domains, we found that all four LLMs achieved higher accuracy on general questions than on clinical questions, although the difference was statistically significant only for Grok-4. Notably, GPT-5, Grok-4, and Gemini 2.5 Pro achieved 99% accuracy on general questions. In clinical questions, Gemini 2.5 Pro still achieved the highest accuracy rate of 97.0%, with GPT-5 and Claude 4.1 Opus also exceeding 95%. Because they generally require factual recall rather than complex reasoning, this near-perfect accuracy further demonstrates that, with training on increasingly large-scale datasets, the latest LLMs have achieved almost textbook-level mastery of fundamental medical knowledge. If ethical and regulatory concerns are addressed, such models can be directly leveraged as educational tools to teach foundational medical knowledge. Nevertheless, improving the performance on clinical questions remains a critical challenge, as these items demand higher-order reasoning and contextual interpretation, highlighting an important direction for the continued advancement of LLMs in medical applications.

After reviewing the explanation of incorrect questions. We found the most common cause for incorrect answers was misunderstanding of the number of selected options. As instructed in the beginning of each section, the default number of correct choices was one. However, LLMs chose more than one in some questions, leading to incorrect results. It is critical to retain the instruction contents while analyzing the questions until the end of each session. The second most common cause for incorrect answers was misdiagnosis from clinical images such as CT or misunderstanding of illustrations. Gemini 2.5 pro performed better in answering questions with images. Nevertheless, improvement in comprehending images, especially those images that directly contributes to a final diagnosis, remains a common challenged shared by LLMs. In addition, LLMs lack the common knowledge in perceiving the directions of clinical images: they always got the left and right directions wrong on X-ray images or CT. Third, although LLMs presented with high performance in collecting evidence from publicly available information such as clinical guidelines and textbooks, they seem to have a long way to go when it comes to prioritizing clinical actions. They tend to choose incorrect options or falsely choose multiple options in questions asking “the most appropriate action as the immediate next step”.

Finally, it is crucial to note that near-perfect performance on medical licensing examinations does not necessarily translate into clinical safety or readiness for autonomous use. Licensing exams primarily test factual knowledge and structured reasoning, whereas real-world clinical practice requires additional competencies such as patient communication, physical examination, contextual judgment, ethical considerations, and accountability for outcomes. Current LLMs cannot replicate these broader dimensions of medical expertise. Therefore, while our findings underscore the potential of advanced LLMs as powerful tools for medical education and as adjuncts to decision support, their safe and responsible integration into clinical workflows requires rigorous real-world validation, careful regulatory oversight, and the continued presence of human clinicians to ensure patient safety.

### 4.2. Limitation

This study exclusively evaluated the performance of LLMs on the JNME, which is written in Japanese. Therefore, these findings may not be generalizable to medical licensing examinations in other countries or languages. However, previous studies indicated that LLMs tend to perform better on examinations written in English than those presented in other languages [4]. Based on this evidence, it is reasonable to hypothesize that the four LLMs tested in the present study might achieve even higher and potentially near-perfect scores on medical examinations delivered in English. In contrast, their performance may be lower in examinations that incorporate traditional medicine domains (e.g., Traditional Chinese Medicine or Korean Traditional Medicine), where relevant training data are likely limited [41].i Therefore, future studies should examine the performance of LLMs across diverse linguistic and cultural contexts to better assess their global applicability in medical education and licensing.

### 4.3. Conclusion

This study evaluated the performance of GPT-5, Grok-4, Claude Opus 4.1, and Gemini 2.5 Pro on the JNME. All four LLMs achieved accuracies exceeding 95%, markedly higher than those reported in previous studies. To the best of our knowledge, this is the first study to demonstrate that LLMs can surpass the 95% accuracy threshold across an entire set of medical licensing examination questions, representing a milestone in their potential application in medical education and clinical decision support.

Although image-based questions, clinical questions, and higher-difficulty questions negatively affected the performance of these models, the magnitude of these effects was substantially smaller than that observed in earlier-generation LLMs. This reflects the enhanced stability, internal consistency, and robustness of the latest models in handling complex question formats and reasoning challenges. In this study, the main patterns of errors in LLMs were selecting extra options and failing to correctly identify left and right during X-ray and CT image recognition.

In particular, Gemini 2.5 Pro achieved the highest overall accuracy (97.2%) and consistently maintained a performance above 95%, even in subgroups traditionally disadvantageous for LLMs (e.g., image-based and clinical questions), demonstrating exceptional robustness and reliability. In contrast, Grok-4 showed more pronounced performance gaps between image-based and non-image-based questions and between general and clinical questions.

## Supporting information

Complete outputs with correctness annotations.

## Data Availability

All data produced in the present work are contained in the manuscript.

## Author Contributions

Conceptualization: Mingxin Liu, Tsuyoshi Okuhara; Methodology: Mingxin Liu, Tsuyoshi Okuhara, Zhehao Dai, Wenqiang Yin; Formal analysis and investigation: Mingxin Liu, Tsuyoshi Okuhara, Wenqiang Yin, Hiroko Okada; Writing – original draft preparation: Mingxin Liu, Tsuyoshi Okuhara; Writing – review and editing: Mingxin Liu, Tsuyoshi Okuhara, Zhehao Dai, Minghong Zhao, Wenqiang Yin, Hiroko Okada, Emi Furukawa, Takahiro Kiuchi; Funding acquisition: Mingxin Liu; Resources: Takahiro Kiuchi; Supervision: Tsuyoshi Okuhara

## Ethics Declarations

### Ethics Approval

Not applicable.

### Content to Participate

Not applicable.

### Clinical trial number

Not applicable.

